# Cognitive profile of mild behavioral impairment in Brain Health Registry participants

**DOI:** 10.1101/2021.07.19.21260787

**Authors:** F. Kassam, H. Chen, R.L. Nosheny, A. McGirr, T. Williams, Nicole Ng, Monica Camacho, R.S. Mackin, M.W. Weiner, Z. Ismail

## Abstract

**INTRODUCTION:** Dementia assessment includes cognitive and behavioral testing with informant validation. Conventional testing is resource intensive, with uneven access. Online unsupervised assessments could reduce barriers to risk assessment. We interrogated the relationship between informant-rated behavioral changes and neuropsychological test performance in older adults in the Brain Health Registry.

**METHODS:** Participants completed online unsupervised cognitive tests, and informants completed the Mild Behavioral Impairment Checklist via a Study Partner portal. Cognitive performance was evaluated in MBI+/- individuals, as was the association between cognitive scores and MBI symptom severity.

**RESULTS:** Mean age of the 499 participants was 67, 61% of which were female. MBI+ participants had lower working memory and executive function test scores. Lower cognitive test scores associated with greater MBI burden.

**DISCUSSION:** Our findings support the feasibility of remote, informant-reported behavioral assessment and support its validity by demonstrating a relationship to cognitive test performance using online unsupervised assessments for dementia risk assessment.

**RESEARCH IN CONTEXT:** *Systematic review:* The authors searched MEDLINE and Google Scholar for studies linking Mild Behavioral Impairment (MBI) and cognition in non-demented older adults. Most studies have utilized transformed Neuropsychiatric Inventory scores to assess MBI, and relatively few using the novel MBI-checklist (MBI-C), with the largest study using self-report. Exploration of informant reports of MBI is important due to impaired insight that may accompany neuropsychiatric symptoms.

*Interpretation:* Older adults with online, informant reported MBI had poorer performance in memory and executive function measured using online neuropsychological testing compared to those without MBI. These findings are consistent with the current literature and suggest that the MBI-C may serve as a marker for poorer cognitive performance.

*Future directions:* Our data support the role of online testing of cognition and behavior for risk assessment. This approach to evaluate behavior and cognition can be explored further, to determine if it is a scalable, online approach to detection of neurodegenerative disease.

## 1. Background

Access to dementia assessments is uneven in North America and across the world. This disparity has important clinical repercussions, particularly in regions where specialized resources are limited, and identification is delayed until later manifestations. The development of unsupervised platforms that do not require highly trained administrators may resolve this dual impasse and improve clinical outcomes. Moreover, they may create a low-cost recruitment infrastructure for early intervention trials, where no disease-modifying drug in Alzheimer’s disease (AD) has met all primary endpoints^1,2^ in part due to poor recruitment of individuals without overt impairment or who are in the earliest stages of disease^3,4^. Advances in online services open the possibility of assessment portals in any region with an internet connection, for any individual with access to the internet and a computing device. Moreover, cognitive tests have been computerized and can be delivered without an administrator, and with convergent validity with those administered in tertiary cognitive assessment centers^5-9^.

The addition of behavioral assessments to online platforms may provide additional relevant information. Neuropsychiatric symptoms (NPS) such as agitation, anxiety, apathy, depression, and psychosis are considered core features of dementia and are associated with poorer patient outcomes^10^. However, NPS can often precede cognitive symptoms, including in 30% of those who develop AD^11^. Mild behavioral impairment (MBI) is a pre-dementia neurobehavioral syndrome characterized by the *de novo* emergence and persistence of NPS in older adults representing a change from longstanding patterns of behavior^12^. MBI is associated with amyloid, tau, and neurodegeneration^13-19^, and a greater risk of incident cognitive decline and dementia^20-26^. Incorporating MBI into screening may provide a complementary approach to early detection^4^. However, informant information is often required to validate the syndrome, and structured assessment tools suitable for widespread dissemination through unsupervised platforms have only recently been developed. The Mild Behavioral Impairment Checklist (MBI-C) incorporates informant information and is the validated case ascertainment instrument developed specifically to capture MBI in accordance with the criteria developed by the International Society to Advance Alzheimer’s Research and Treatment-Alzheimer’s Association (ISTAART-AA)^27-30^. Translated into over 20 languages, the MBI-C may also allow a broader reach for obtaining online informant reports of behavioral change.

The aim of this study was to investigate informant-based MBI in an online unsupervised platform, the Brain Health Registry (BHR), capable of assessing early dementia risk markers^31^. We determined the utility of the BHR for converging assessments of cognitive and behavioral symptoms using neuropsychological testing and informant-reported MBI-C. We hypothesized that participants with MBI+ status would have poorer cognitive performance measured by the Lumos test battery. We further hypothesized that individuals with poorer memory, executive function, processing speed, and inhibitory control would have a higher burden of MBI symptoms.

## 2. Methods

### 2.1 Brain Health Registry

The BHR^31^ is an internet-based public registry and cohort that recruits participants using a variety of methods including a website, social media, brochures, and online advertising. All participants are required to give informed consent with an online consent form. Upon completion of the consent form, participants may complete questionnaires regarding personal and family medical history, early childhood history, sleep quality, diet, quality of life scales, psychiatric symptomatology, as well as online cognitive testing via Lumosity^32^, CogState^33^, or Memtrax^34^ tests. Additionally, study partners of BHR participants can register on the BHR study partner portal, on which informant-rated measures are completed^35^.

### 2.2 Study Participants

Participants were included if: 1) Lumosity cognitive tests were completed; and 2) their informant completed the MBI-C via the BHR study partner portal within a year of the cognitive tests. Participants were excluded if they reported: 1) developmental or learning disorders; 2) neurological conditions such as movement disorders, multiple sclerosis, traumatic brain injury; 3) current or past psychiatric diagnoses including schizophrenia, major mood or anxiety disorders, or PTSD.

### 2.3 Study Variables

#### 2.3.1 Lumosity online Forward Memory Span

The assessment of Forward Memory Span is based on the Corsi block-tapping tasks^36^. The participant is asked to recall the sequence of circles in the same order it was presented. The length of the sequence increases by one every two trials. The session comes to an end when the participant records two incorrect answers at the same span level. This task is used as a measure of visual short-term memory and attention.

#### 2.3.2 Lumosity online Reverse Memory Span

The Reverse Memory Span task is a slightly altered version of the original Corsi block-tapping tasks. It is identical to the forward visual memory span assessment, with the exception that the participant is asked to recall the sequence of circles in the reverse order. This reverse task is used as a measure of visual working memory and attention.

#### 2.3.3 Lumosity online Trail Making Test B

In Trail Making Test (TMT) B, blue circles (numbered 1 to 12) and capital letters (A to L) are arranged in 6 possible layouts with non-overlapping spatial locations. The participant must alternate between numbers and letters for this task, clicking in increasing order. When the blue circle is clicked, it turns orange and a straight line appears to connect the circles. The timer for the task begins when the participants clicks the first circle. If the participant records an incorrect click, an X appears on their screen and they are required to go back to the previous circle. For this study, we included the response time and number of errors as measures of processing speed attention and sequencing ability.

#### 2.3.4 Lumosity online Go/No-Go

In the Go/No-Go assessment, participants are presented with target pictures and distractor stimuli. The target picture is chosen from a set of photos of fruit. Each stimulus appears after a random delay between 1000 and 3000 ms to discourage anticipatory responding. The participant is instructed to respond as quickly as possible within 1500ms. The assessment ends when a participant responds to ten “Go” trials. If the participant submits three incorrect responses (responding to “no-go” or failing to respond to “go”), the participant will restart the task. The participant is given feedback on timing and correctness. This assessment is used to measure response inhibition and speed of information processing.

#### 2.3.5 Mild Behavioral Impairment Checklist

The MBI-C is included in the BHR study partner portal and is therefore completed by an informant. The MBI-C is explicit that symptoms are *de novo* in later life, represent a change from longstanding patterns of behavior, and are persistent for at least six months. The MBI-C consists of questions in the five MBI domains of apathy, mood and anxiety, agitation and impulsivity, impaired social cognition, and psychosis, with items geared towards capturing NPS in community dwelling, functionally independent, non-demented older adults. The scale takes ∼7-8 minutes to complete, consisting of 34 questions; scoring is from 0-3, representing absent, mild, moderate, and severe changes, with a total score range of 0-102^28^.

### 2.4 Statistical Analysis

Continuous demographic variables (age and years of education) were analyzed using independent sample t-tests to compare MBI+ and MBI-groups; sex distribution between the two groups was analyzed using chi-square tests. MBI-C was dichotomized based on a validation in primary care non-demented older adults in which scores of >7 differentiated MBI+ from MBI-with a sensitivity of 0.93, specificity of 0.76 and AUC of 0.93^37^. As exploratory analyses, cutpoints of >5 and >6 were also analyzed. Univariate Analysis of Covariance (ANCOVA) was used to compare performance on Lumosity cognitive tests between MBI+ and MBI-groups, covarying for age, sex, education, and neuropsychological and neurobehavioral assessment interval. Skewed data were log-transformed, however the TMT response time variable was analyzed with a negative binomial regression due a skewed distribution with an overrepresentation of zeros. For Go/No-Go errors, ordinal logistic regression was performed because the response variable only had three possible values: 0, 1, and 2.

Additionally, negative binomial regressions were fitted to assess Lumosity task prediction of MBI-C total scores. Negative binomial regression is preferred when the data are skewed, as in this sample where the mode on the MBI-C is zero indicating no emergent and persistent NPS. Lumosity task measures as continuous scores were the independent variables in these models. The covariates included were age, sex, education, and neuropsychological and neurobehavioral assessment interval. The *p* values for Lumosity task measures were calculated using likelihood ratio tests.

Statistical analyses were performed using SPSS v26 and R 3.6.2.

## 3 Results

Participant selection is described in Figure 1. The final sample included 499 participants with a mean age of 67 (SD 10.4), of which 308/499 were females (61%) (Table 1). The number of MBI+ participants was 31 (6.2%) (Figure 1). A significantly greater number of men were classified as MBI+ (64%, p=0.002). MBI+ participants had significantly poorer forward memory span (mean sequence length of 4.68 vs. 5.26, p=0.005; Figure 2a), poorer reverse memory span (3.81 vs. 4.85, p<0.0001; Figure 2b), more TMT errors (4.29 vs. 1.85, p=0.01; Figure 2c), and longer TMT completion time (67.67 vs. 45.08 seconds, p<0.0001; Figure 2d). MBI was not associated with the number of errors (p=0.84) and response time (p=0.16) on the Go/No-Go task (Figures 2e & 2f). The effect sizes for these differences were modest. Of the tests that significantly differed between groups, the largest effect size (Cohen’s *f*) was for reverse memory span (0.20), followed by TMT response time (0.20), memory span (0.13), and TMT accuracy (0.11). See Tables 2-4 for statistical reporting. Analyses using cutpoints of >5 and >6 for MBI-C show very similar results and are included in supplemental tables (Supplemental tables 1-6).

**Table 1.** Summary statistics for demographics.

| *Cutpoint of MBI-C > 7* |  |  |  |  |
| --- | --- | --- | --- | --- |
|  | MBI+ (n=31) | MBI- (n=468) | Test Statistic | *p* |
| Average Age | 69.52 | 67.11 | $t(497)=1.25$ | .213 |
| Average Estimated Years of Education | 17.58 | 17.14 | $t(497)=1.13$ | .260 |
| Number of Females | 11 | 297 |  |  |
| Percentage of Female | 35.48% | 63.46% | $\chi^2=9.63$ | **.002** |

**Table 2.** Summary statistics for Lumosity tasks (ANCOVA)

| Cognitive Measures | Mean (MBI+) | Mean (MBI-) | *F* | *df* within | *df* between | Partial $\eta^2$ (effect size) | *P* Value |
| --- | --- | --- | --- | --- | --- | --- | --- |
| *Cutpoint of MBI-C > 7* |  |  |  |  |  |  |  |
| Memory Span | 4.68 | 5.26 | 8.11 | 1 | 493 | 0.016 (0.13) | **0.0046** |
| Reverse Memory Span | 3.81 | 4.85 | 20.06 | 1 | 492 | 0.039 (0.20) | **< 0.0001** |
| Trailmaking Errors | 4.29 | 1.85 | 6.81 | 1 | 493 | 0.014 (0.11) | **0.0093** |
| Trailmaking Response Time (log transformed) | 67.67 | 45.08 | 19.40 | 1 | 493 | 0.038 (0.20) | **< 0.0001** |
| Go/No-Go Errors | 0.71 | 0.64 | 0.04 | 1 | 491 | 0.000 (0.01) | 0.8386 |
| Go/No-Go Response Time | 496.42 | 473.87 | 1.98 | 1 | 491 | 0.004 (0.06) | 0.1604 |

**Table 3.**
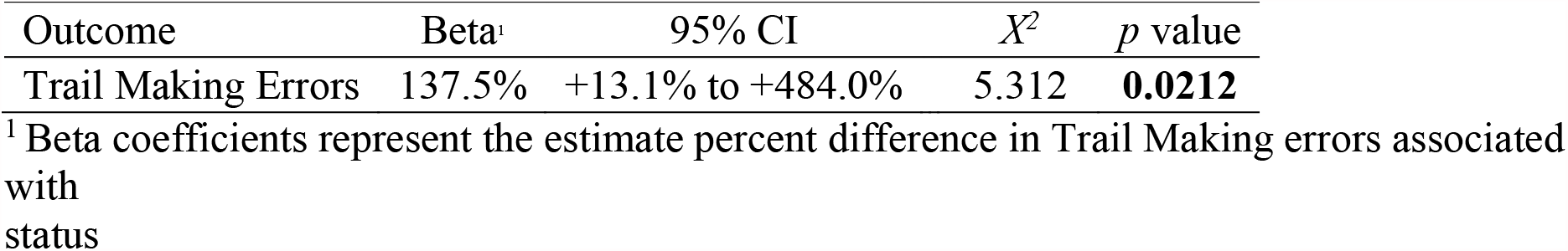
Summary statistics for using MBI-C status (Cutpoint of 7) to predict Trail Making Errors (Negative Binomial Regression)

**Table 4.** Summary statistics for using MBI-C status (Cutpoint of 7) to predict Go/No-Go Errors (Ordinal Logistic Regression)

| Outcome | Odds Ratio | 95% CI | *p* value |
| --- | --- | --- | --- |
| GoNoGo Errors | 1.058 | 0.517 to 2.121 | 0.875 |

**Figure 1.**
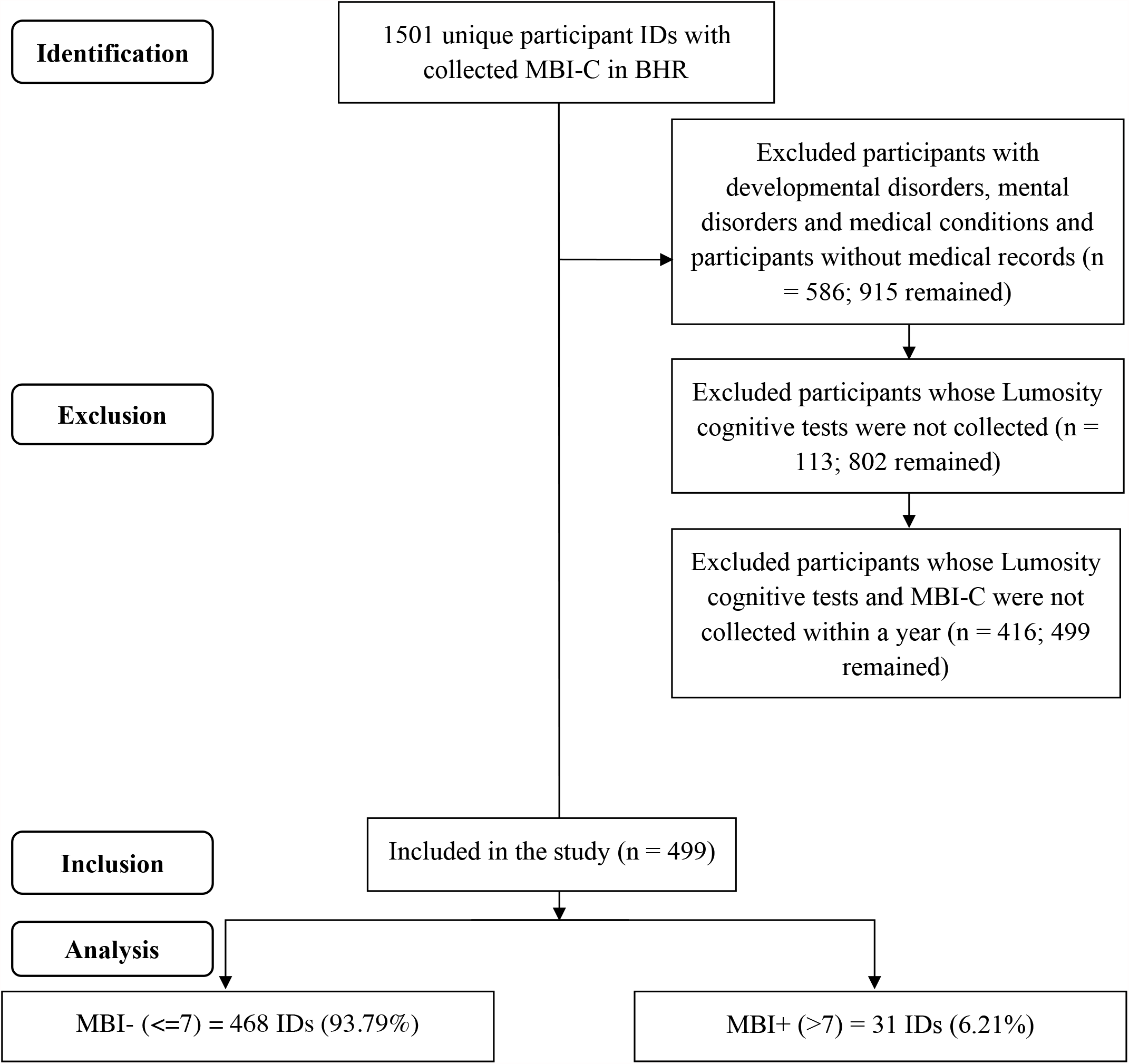
Flowchart of participants from the BHR included for analysis.

**Figure 2a.**
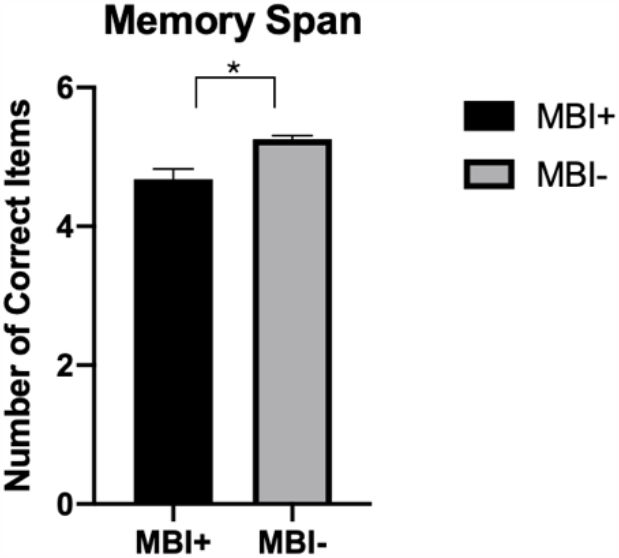
Positive result (>7) on the MBI-C is associated with shorter memory span

**Figure 2b.**
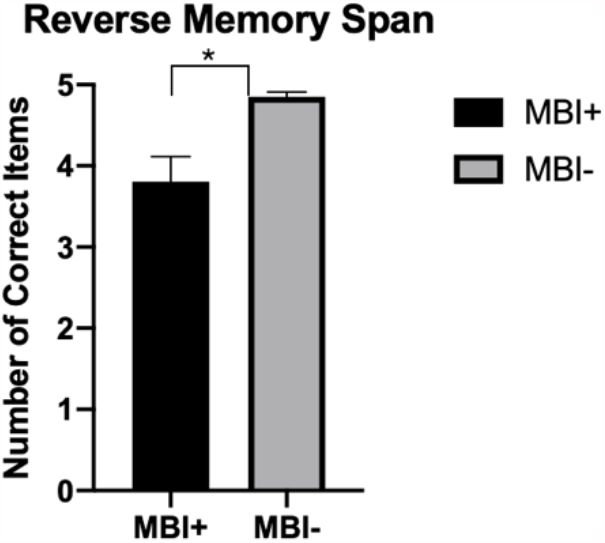
Positive result (>7) on the MBI-C is associated with shorter reverse memory span

**Figure 2c.**
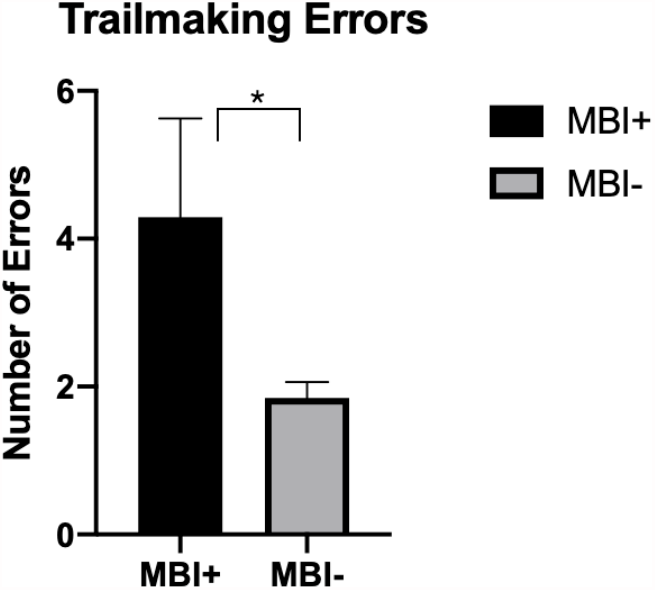
Positive result (>7) on the MBI-C is associated with more errors in the Trailmaking-B task

**Figure 2d.**
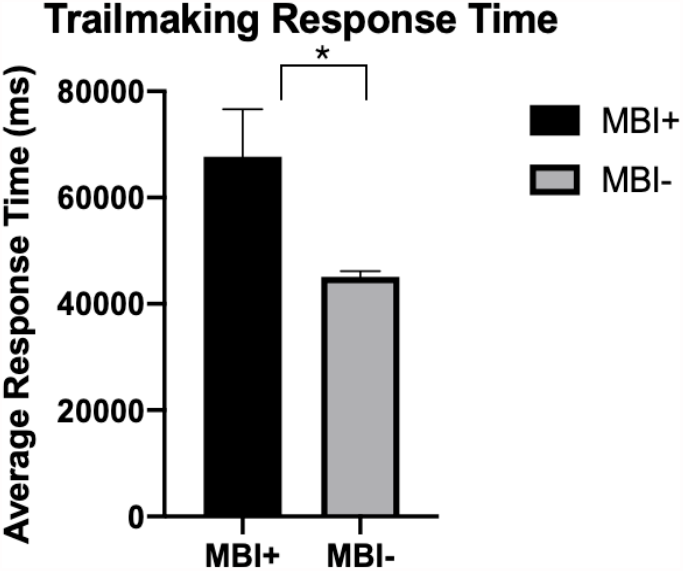
Positive result (>7) on the MBI-C is associated with longer response time in Trailmaking-B task

**Figure 2e.**
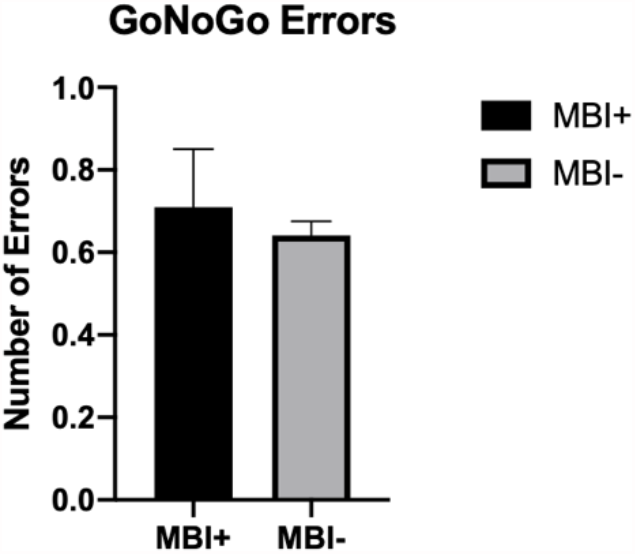
Positive result (>7) on the MBI-C is not associated with the number of errors on a GoNoGo task

**Figure 2f.**
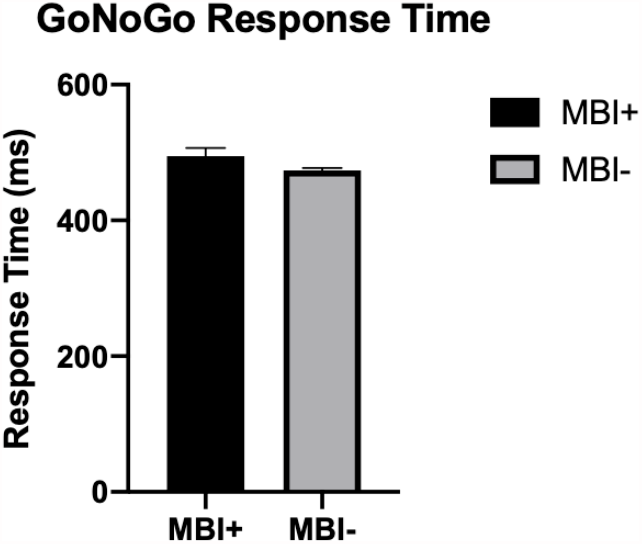
Positive result (>7) on the MBI-C is not associated with the response time on a GoNoGo task

Negative binomial regressions utilizing Lumosity scores to predict MBI score determined that worse memory span (*X*^*2*^(1, N=499)=6.6, p=0.01), worse reverse memory span *X*^*2*^(1, N=498)=5.4, (p= 0.02), more TMT errors (*X*^*2*^(1, N=499)=5.8, p=0.02) and longer TMT response time (*X*^*2*^(1, N=499)=9.6, p=0.002) were all associated with higher MBI-C total scores. Go/No-Go errors (*X*^*2*^(1, N=497)=0.16, p=0.69) and Go/No-Go response time (*X*^*2*^(1, N=497)=0.97, p=0.33) were not associated with MBI-C score (Table 5).

**Table 5.** Summary statistics for Lumosity tasks predicting MBI-C total score.

| Predictor | Beta<sup>1</sup> | 95% CI | $\chi^2$ | *p* value |
| --- | --- | --- | --- | --- |
| Memory Span | -26.2% | -41.8% to -7.1% | 6.6513 | **0.0099** |
| Reverse Memory Span | -15.7% | -27.7% to -2.6% | 5.3765 | **0.0204** |
| Trail Making Errors | 5.3% | +0.9% to +11.2% | 5.8059 | **0.0160** |
| Trail Making Response Time | 0.0012% | +0.0004% to +0.0023% | 9.6268 | **0.0019** |
| GoNoGo Errors | -6.2% | -31.0% to +29.0% | 0.1594 | 0.6897 |
| GoNoGo Response Time | 0.1% | -0.2% to +0.5% | 0.9678 | 0.3252 |

<sup>1</sup> Beta coefficients represent the estimate percent difference in total MBI-C score given one unit change in Lumosity task measure.

## 4 Discussion

In a sample of 499 participant dyads in BHR, we demonstrated the feasibility of delivering unsupervised online assessments of behavioral and cognitive markers of dementia risk. Utilizing the validated cut off score of >7 on the MBI-C, MBI+ status was associated with significant differences in memory and executive function, measured using memory span, reverse memory span, TMT errors and TMT speed. Further, significant associations were found between poorer objectively measured cognitive performance, in the domains of memory and executive function, and MBI symptom severity. Effect sizes were small, ranging from 0.11-0.20. The findings do suggest that a simple informant reported behavioral measure completed via an online portal might be a relevant addition to neuropsychological testing, warranting further study in BHR. In other work, MBI+ status has demonstrated significant and meaningful associations with incident cognitive decline and dementia across several studies, settings, and populations^20-26^. Thus, while convergent with tests of memory and executive function, behavioral and cognitive markers of risk may be distinct, potentially offering complementary measures of risk.

Our data indicate that the BHR is an effective platform to conduct remote assessments of cognitive functioning with convergence of behavioral and cognitive tests. Poorer performance on unsupervised online neuropsychological testing has been associated with self-report MCI and AD^8^. Online participant testing is an efficient and reliable tool for neuropsychological testing, which can identify performance decrements in executive dysfunction and memory^32^. Similarly, online informant reports such as those collected in the BHR study partner portal are valuable and informative. Online study-partner reported cognitive decline is comparable to data collected in clinic, is associated with objectively defined participant cognition^35^, and is associated with amyloid, clinical diagnosis of dementia due to AD, and in-clinic cognitive screening test scores^5^. In our study, online informant-reported behavioral symptoms associated with differences in memory and executive function collected via the participant portal. This harmonized utilization of participant and study partner portals is effective and can allow continuation of research activities even during trying times such as the recent pandemic, where consistent in person visits between clinicians and patients were not feasible. Although the COVID-19 pandemic has highlighted the need for alternative infrastructure to allow continued care, the tools that have been developed may permit the assessment of older adults who for physical, social, or cognitive reasons could not previously access care. This approach also allows outreach to areas less accessible to academic centers.

The cognitive domain differences detected with the MBI-C include memory and executive function, which are relevant and important for AD risk^38^. Early decline in memory and executive function has been shown to be associated with the preclinical disease process, thus, early detection of reductions in cognitive functioning may be useful in identifying populations at risk^39,40^. Both memory and executive function are important endpoints in AD trials^41^. Longitudinal cohorts have demonstrated that an acceleration of decline in memory performance occurs 3-4 years before a diagnosis of MCI and 7 years before a diagnosis of AD, while for executive function an accelerated decline occurs 2-3 years before AD diagnosis^42,43^. The finding of small but significant associations between MBI and poorer memory and executive function performance is consistent with the evolving description of the cognitive profile of MBI^44-46^. These findings are consistent with a previous study using the UK online PROTECT study portal in which self-reported MBI, measured with the MBI-C, was associated with faster decline in attention and working memory at one year in older adults with normal cognition^20^. A subsequent analysis of cognitively normal PROTECT participants, with a median follow up time of 3 years, demonstrated an association between baseline informant-reported MBI+ status and decline in measures of working memory and fluid intelligence^26^. In an overlapping sample, AD genetic risk was determined using polygenic risk scores. AD genetic risk was associated with worse cognition in the informant-reported MBI+ group but not in the MBI-group. The strongest association was in those with more severe MBI, aged ≥65^47^. These convergent findings support leveraging online cognitive and behavioral measures to explore dementia risk. In a recent study of National Alzheimer Coordinating Center data, the combination of informant-reported MBI and subjective cognitive decline (SCD) had a greater risk of incident cognitive and functional decline at three years compared to either MBI or SCD alone^24^. Further, in a subsequent study, those with SCD and MBI together had a shorter median time to incident MCI compared to those with SCD in the absence of MBI^48^. Taken together the results suggest that individuals with subtle neuropsychological test score differences and MBI together may be at higher risk for cognitive and functional decline.

As the case ascertainment instrument developed to measure MBI in accordance with the ISTAART-AA MBI criteria, the MBI-C was designed to: 1) operationalize the MBI concept; 2) measure a selected list of NPS which may help identify preclinical or prodromal dementia; and 3) predict risk of several dementias^28^. This instrument has been validated in an online cohort of cognitively normal older adults^27^, a primary care sample with SCD^29^ or MCI^49^, and a cognitive neurology clinic population with SCD and MCI^50^.

However, limitations in our study may affect interpretation and generalizability. These limitations include high education levels and restriction to participants and study-partners who can successfully complete tasks online^35^. Since the BHR is an online self-report database, the lack of a clinical diagnosis within the sample group is a potential source of error. BHR participants may have undiagnosed and/or unreported neurodegenerative disease or psychiatric disorders, which may be associated with greater MBI score. While online cognitive testing has been validated^8,33^, further research is needed, given the lack of supervision or control for test environment, external factors, distractors, or cues. Further, we were not able to control for important disease related factors such as severity and time since symptom onset. Although MBI was associated with statistically significant differences in Lumosity neuropsychological test scores, effect sizes were small, and the clinical significance of these differences is difficult to interpret in the largely cognitively healthy population enrolled in BHR. Our data are promising but not conclusive, and further research is required. Whether these small differences in cognitive test scores represent greater risk for incident cognitive decline and dementia can be addressed in the future using longitudinal data and a cohort that includes participants with cognitive impairment.

In summary, in this BHR study combining self- and informant-rated measures, we observed the convergence of behavioral risk markers for dementia and cognitive differences, reflected by neuropsychological tests incorporating memory and executive function. The findings lend additional support to online unsupervised administration of cognitive and neuropsychiatric measures, as a low-cost approach to improve access to neurocognitive assessments, potentially identifying at-risk older adults.

## Data Availability

Data used in the manuscript is used from the Brain Health Registry online platform.

## Acknowledgements

Data used in this study were collected using the BHR, which is funded by the NIH, Alzheimer’s Association, Alzheimer’s Drug Discovery Foundation, California Department of Public Health, Connie and Kevin Shanahan, The Drew Foundation, General Electric, Global Alzheimer’s Platform Foundation, Larry L. Hillbolm Foundation, The Ray and Dagmar Dolby Family Fund, The Rosenberg Alzheimer’s Project and Patient-Centered Outcomes Research Institute.

FK, HC, AW, TW have no interests to declare. ZI is funded by the Canadian Institutes of Health Research, and has received consulting fees/honoraria from Otsuka/Lundbeck, outside the submitted work. His institution has received funds from Acadia, Biogen, Roche, and Sunovion, also outside the submitted work. RLN is a co-investigator for the BHR. RSM has received grant funding from the National Institute of Mental Health and has received research support from Johnson & Johnson. MWW receives support for his work from the following: National Institute of Health, Department of Defense, Patient-Centered Outcomes Research Institute, California Department of Public Health, University of Michigan, Siemens, Biogen, Larry L. Hillbolm Foundation, Alzheimer’s Association, The State of California, Johnson & Johnson, Kevin and Connie Shanahan, GE, Vrije Universiteit Medical Center Amsterdam, Australian Catholic University, The Stroke Foundation and the Veterans Administration. He has served on the Advisory Boards for Eli Lilly, Cerecin/Accera, Roche, Alzheon, Inc., and BHR.

## HIGHLIGHTS

- Online portals for older adults and study partners can be used for dementia risk assessment
- This approach is useful when in-person assessments are not feasible (physical, social, cognitive or health systems-realted (e.g., the COVID-19 pandemic))
- The relationship between mild behavioral impairment (MBI) and cognition in older adults was explored
- MBI was associated with small magnitude, but significantly poorer performance in memory and executive function, and may serve as a complementary measure of risk

## References

1. Marsden G, Mestre-Ferrandiz J. Dementia: The R&D Landscape. 2015.

2. Cummings J, Reiber C, Kumar P. The price of progress: Funding and financing Alzheimer’s disease drug development. Alzheimer’s & Dementia: Translational Research & Clinical Interventions. 2018;4:330–343.

3. Gauthier S, Albert M, Fox N, et al. Why has therapy development for dementia failed in the last two decades? Alzheimers & Dementia. 2016;12(1):60–64.

4. Mortby ME, Black SE, Gauthier S, et al. Dementia clinical trial implications of Mild Behavioral Impairment. Int Psychogeriatr. 2018;30(2):171–175.

5. Nosheny RL, Camacho MR, Jin C, et al. Validation of online functional measures in cognitively impaired older adults. Alzheimer’s & Dementia. 2020;16(10):1426–1437.

6. Brooker H, Williams G, Hampshire A, et al. FLAME: A computerized neuropsychological composite for trials in early dementia. Alzheimers Dement (Amst). 2020;12(1):e12098.

7. Papp KV, Rentz DM, Maruff P, et al. The computerized cognitive composite (c3) in a4, an alzheimer’s disease secondary prevention trial. The journal of prevention of Alzheimer’s disease. 2021;8(1):59–67.

8. Mackin RS, Insel PS, Truran D, et al. Unsupervised online neuropsychological test performance for individuals with mild cognitive impairment and dementia: results from the Brain Health Registry. Alzheimers Dement (Amst). 2018;10:573–582.

9. Perin S, Buckley RF, Pase MP, et al. Unsupervised assessment of cognition in the Healthy Brain Project: Implications for web-based registries of individuals at risk for Alzheimer’s disease. Alzheimer’s & Dementia: Translational Research & Clinical Interventions. 2020;6(1):e12043.

10. Lanctôt KL, Amatniek J, Ancoli-Israel S, et al. Neuropsychiatric signs and symptoms of Alzheimer’s disease: New treatment paradigms. Alzheimer’s & Dementia: Translational Research & Clinical Interventions. 2017;3:440–449.

11. Wise EA, Rosenberg PB, Lyketsos CG, Leoutsakos J-M. Time course of neuropsychiatric symptoms and cognitive diagnosis in National Alzheimer’s Coordinating Centers volunteers. Alzheimers Dement (Amst). 2019;11:333–339.

12. Ismail Z, Smith EE, Geda Y, et al. Neuropsychiatric symptoms as early manifestations of emergent dementia: provisional diagnostic criteria for mild behavioral impairment. Alzheimer’s & Dementia. 2016;12(2):195–202.

13. Andrews SJ, Ismail Z, Anstey KJ, Mortby M. Association of Alzheimer’s genetic loci with mild behavioral impairment. Am J Med Genet B Neuropsychiatr Genet. 2018;177(8):727–735.

14. Creese B, Brooker H, Aarsland D, Corbett A, Ballard C, Ismail Z. Genetic risk for Alzheimer disease, cognition and Mild Behavioral Impairment in healthy older adults. Alzheimer’s & Dementia: DADM. 2021;in press:2020.2005.2013.20100800.

15. Johansson M, Stomrud E, Insel P, et al. Mild Behavioral Impairment and its relation to tau pathology in preclinical Alzheimer’s disease. Transl Psychiatry. 2021;11(76).

16. Lussier FZ, Pascoal TA, Chamoun M, et al. Mild behavioral impairment is associated with β-amyloid but not tau or neurodegeneration in cognitively intact elderly individuals. Alzheimer’s & Dementia. 2020;16:192–199.

17. Naude J, Gill S, Hu S, et al. Plasma Neurofilament Light: a marker of cognitive decline in Mild Behavioural Impairment. J Alzheimers Dis. 2020;76(3):1017–1027.

18. Gill S, Wang M, Mouches P, et al. Neural Correlates of the Impulse Dyscontrol Domain of Mild Behavioral Impairment. Int J Geriatr Psychiatry. 2021;in press.

19. Matuskova V, Ismail Z, Nikolai T, et al. Mild behavioral impairment is associated with atrophy of entorhinal cortex and hippocampus in a memory clinic cohort. Frontiers in Aging Neuroscience. 2021;13:236.

20. Creese B, Brooker H, Ismail Z, et al. Mild Behavioral Impairment as a Marker of Cognitive Decline in Cognitively Normal Older Adults. The American Journal of Geriatric Psychiatry. 2019;27(8):823–834.

21. Gill S, Mouches P, Hu S, et al. Using Machine Learning to Predict Dementia from Neuropsychiatric Symptom and Neuroimaging Data. J Alzheimers Dis. 2020;75(1):277– 288.

22. Matsuoka T, Ismail Z, Narumoto J. Prevalence of mild behavioral impairment and risk of dementia in a psychiatric outpatient clinic. J Alzheimers Dis. 2019;70(2):505–513.

23. Taragano FE, Allegri RF, Heisecke SL, et al. Risk of Conversion to Dementia in a Mild Behavioral Impairment Group Compared to a Psychiatric Group and to a Mild Cognitive Impairment Group. J Alzheimers Dis. 2018;62:227–238.

24. Ismail Z, McGirr A, Gill S, Hu S, Forkert ND, Smith EE. Mild Behavioral Impairment and Subjective Cognitive Decline predict Cognitive and Functional Decline. J Alzheimers Dis. 2021;80:459–469.

25. Tsunoda K, Yamashita T, Osakada Y, et al. Positive baseline behavioral and psychological symptoms of dementia predict a subsequent cognitive impairment in cognitively normal population. Neurology and Clinical Neuroscience. 2021.

26. Wolfova K, Creese B, Aarsland D, et al. Sex differences in the association of mild behavioral impairment with cognitive aging. medRxiv. 2021.

27. Creese B, Griffiths A, Brooker H, et al. Profile of mild behavioral impairment and factor structure of the mild behavioral impairment checklist in cognitively normal older adults. Int Psychogeriatr. 2020;32(6):705–717.

28. Ismail Z, Aguera-Ortiz L, Brodaty H, et al. The Mild Behavioral Impairment Checklist (MBI-C): A Rating Scale for Neuropsychiatric Symptoms in Pre-Dementia Populations. Journal of Alzheimers Disease. 2017;56(3):929–938.

29. Mallo SC, Ismail Z, Pereiro AX, et al. Assessing mild behavioral impairment with the mild behavioral impairment checklist in people with subjective cognitive decline. Int Psychogeriatr. 2019;31(2):231–239.

30. Saari T, Smith EE, Ismail Z. Network analysis of impulse dyscontrol in mild cognitive impairment and subjective cognitive decline. Int Psychogeriatr. 2021:1–10.

31. Weiner MW, Nosheny R, Camacho M, et al. The Brain Health Registry: An internet-based platform for recruitment, assessment, and longitudinal monitoring of participants for neuroscience studies. Alzheimer’s & Dementia. 2018;14(8):1063–1076.

32. Morrison GE, Simone CM, Ng NF, Hardy JL. Reliability and validity of the NeuroCognitive Performance Test, a web-based neuropsychological assessment. Front Psychol. 2015;6:1652.

33. Lim YY, Pietrzak RH, Bourgeat P, et al. Relationships between performance on the Cogstate Brief Battery, neurodegeneration, and Aβ accumulation in cognitively normal older adults and adults with MCI. Arch Clin Neuropsychol. 2015;30(1):49–58.

34. Ashford JW, Gere E, Bayley PJ. Measuring memory in large group settings using a continuous recognition test. J Alzheimers Dis. 2011;27(4):885–895.

35. Nosheny RL, Camacho MR, Insel PS, et al. Online study partner-reported cognitive decline in the Brain Health Registry. Alzheimer’s & Dementia: Translational Research & Clinical Interventions. 2018;4:565–574.

36. Milner B. Interhemispheric differences in the localization of psychological processes in man. Br Med Bull. 1971.

37. Mallo SC, Pereiro AX, Ismail Z, et al. Mild Behavioral Impairment Checklist (MBI-C): A Preliminary Validation Study. Alzheimers Dement. 2018;14(7 supplement).

38. Wilson RS, Leurgans SE, Boyle PA, Bennett DA. Cognitive decline in prodromal Alzheimer disease and mild cognitive impairment. Arch Neurol. 2011;68(3):351–356.

39. Nagata T, Shinagawa S, Ochiai Y, et al. Association between executive dysfunction and hippocampal volume in Alzheimer’s disease. Int Psychogeriatr. 2011;23(5):764–771.

40. Almkvist O, Basun H, Bäckman L, et al. Mild cognitive impairment—an early stage of Alzheimer’s disease? In: Alzheimer’s Disease—From Basic Research to Clinical Applications. Springer; 1998:21–29.

41. Vellas B, Andrieu S, Sampaio C, Coley N, Wilcock G. Endpoints for trials in Alzheimer’s disease: a European task force consensus. The Lancet Neurology. 2008;7(5):436–450.

42. Mistridis P, Krumm S, Monsch AU, Berres M, Taylor KI. The 12 years preceding mild cognitive impairment due to Alzheimer’s disease: the temporal emergence of cognitive decline. J Alzheimers Dis. 2015;48(4):1095–1107.

43. Grober E, Hall CB, Lipton RB, Zonderman AB, Resnick SM, Kawas C. Memory impairment, executive dysfunction, and intellectual decline in preclinical Alzheimer’s disease. J Int Neuropsychol Soc. 2008;14(2):266–278.

44. Rouse HJ, Small BJ, Schinka JA, Loewenstein DA, Duara R, Potter H. Mild behavioral impairment as a predictor of cognitive functioning in older adults. Int Psychogeriatr. 2020:1–9.

45. Yoon E, Ismail Z, Hanganu A, et al. Mild Behavioral Impairment is linked to worse cognition and brain atrophy in Parkinson’s disease. Neurology. 2019;93(8):e766–e777.

46. Wong F, Ng KP, Yatawara C, Low A, Ismail Z, Kandiah N. Characterising mild behavioural impairment in Asian mild cognitive impairment and cognitively normal individuals: Neuropsychiatry and behavioral neurology: Neuropsychiatric symptoms in MCI and dementia. Alzheimer’s & Dementia. 2020;16:e045059.

47. Creese B, Arathimos R, Brooker H, et al. Genetic risk for Alzheimer’s disease, cognition, and mild behavioral impairment in healthy older adults. Alzheimers Dement (Amst). 2021;13(1):e12164.

48. Nathan S, Gill S, Ismail Z. APOE ε4 status in pre-dementia risk states, mild behavioural impairment and subjective cognitive decline, and the risk of incident cognitive decline. Paper presented at: 2020 Alzheimer’s Association International Conference 2020.

49. Mallo SC, Ismail Z, Pereiro AX, et al. Assessing mild behavioral impairment with the Mild behavioral impairment-checklist in people with mild cognitive impairment. J Alzheimers Dis. 2018;66(1):83–95.

50. Hu S, Patten SB, Fick G, Smith EE, Ismail Z. VALIDATION OF THE MILD BEHAVIORAL IMPAIRMENT CHECKLIST (MBI-C) IN A CLINIC-BASED SAMPLE. Alzheimer’s & Dementia: The Journal of the Alzheimer’s Association. 2019;15(7):P365.

