## Supplemental Tables 1-6 for "Cognitive profile of mild behavioral impairment in Brain Health Registry participants"

**Supplemental table 1. Summary statistics for demographics at cutpoint 5 and 6**

| ***Cutpoint of MBI-C > 5*** |  |  |  |  |
| --- | --- | --- | --- | --- |
|  | **MBI+ (n=41)** | **MBI- (n=458)** | **Test Statistic** | ***p*** |
| Average Age | 68.30 | 67.16 | *t*(497) = 0.67 | .503 |
| Average Estimated Years of Education | 17.36 | 17.15 | *t*(497) =0 .63 | .529 |
| Number of Females | 19 | 22 |  |  |
| Percentage of Female | 46.34% | 63.10% | *χ*^2^ = 4.47 | **.034** |
| ***Cutpoint of MBI-C > 6*** |  |  |  |  |
|  | **MBI+ (n=34)** | **MBI- (n=465)** | **Test Statistic** | ***p*** |
| Average Age | 69.22 | 67.11 | *t*(497) = 1.14 | .255 |
| Average Estimated Years of Education | 17.24 | 17.16 | *t*(497) = 0.19 | .844 |
| Number of Females | 14 | 294 |  |  |
| Percentage of Female | 41.18% | 63.23% | *χ*^2^ = 6.52 | **.011** |

**Supplemental table 2. Summary statistics for Lumosity tasks (ANCOVA) at cutpoints 5 and 6.**

| **Cognitive Measures** | **Mean (MBI+)** | **Mean (MBI-)** | ***F*** | ***df within*** | ***df between*** | ***Partial η^2^ (effect size)*** | ***P Value*** |
| --- | --- | --- | --- | --- | --- | --- | --- |
| ***Cutpoint of MBI-C > 5*** |  |  |  |  |  |  |  |
| Memory Span | 4.76 | 5.26 | 8.61 | 1 | 493 | 0.017 (0.13) | **0.0035** |
| Reverse Memory Span | 4.12 | 4.84 | 12.76 | 1 | 492 | 0.025 (0.16) | **0.0004** |
| Trailmaking Errors | 3.90 | 1.83 | 6.42 | 1 | 493 | 0.013 (0.11) | **0.0116** |
| Trailmaking Response Time (log transformed) | 65.31 | 44.80 | 20.74 | 1 | 493 | 0.040 (0.21) | **< 0.0001** |
| GoNoGo Errors | 0.66 | 0.64 | 0.01 | 1 | 491 | 0.000 (0.00) | 0.9332 |
| GoNoGo Response Time | 486.39 | 474.28 | 0.79 | 1 | 491 | 0.000 (0.04) | 0.3740 |
| ***Cutpoint of MBI-C > 6*** |  |  |  |  |  |  |  |
| Memory Span | 4.68 | 5.26 | 8.83 | 1 | 493 | 0.018 (0.13) | **0.0031** |
| Reverse Memory Span | 3.88 | 4.85 | 18.11 | 1 | 492 | 0.035 (0.19) | **< 0.0001** |
| Trailmaking Errors | 4.03 | 1.80 | 5.29 | 1 | 493 | 0.011 (0.11) | **0.0218** |
| Trailmaking Response Time (log transformed) | 66.21 | 45.04 | 17.95 | 1 | 493 | 0.035 (0.19) | **< 0.0001** |
| GoNoGo Errors | 0.68 | 0.64 | 0.00 | 1 | 491 | 0.000 (0.00) | 0.9930 |
| GoNoGo Response Time | 494.85 | 473.84 | 1.75 | 1 | 491 | 0.004 (0.06) | 0.1866 |

**Supplemental table 3. Summary statistics for using MBI-C status (Cutpoint of 5) to predict Lumosity tasks (Negative Binomial Regression)**

| Outcome | Beta^1^ | 95% CI | *X^2^* |  | *p* value |
| --- | --- | --- | --- | --- | --- |
| Trailmaking Errors | 110.2% | +8.8% to +356.7% | 4.922 |  | **0.0265** |

^1^ Beta coefficients represent the estimate percent difference in trailmaking errors associated with
status

**Supplemental table 4. Summary statistics for using MBI-C status (Cutpoint of 5) to predict Lumosity tasks (Ordinal Logistic Regression)**

| Outcome | Odds Ratio | 95% CI | *p* value |
| --- | --- | --- | --- |
| GoNoGo Errors | 0.967 | 0.515 to 1.782 | 0.915 |

**Supplemental table 5. Summary statistics for using MBI-C status (Cutpoint of 6) to predict Lumosity tasks (Negative Binomial Regression)**

| Outcome | Beta^1^ | 95% CI | *X^2^* |  | *p* value |
| --- | --- | --- | --- | --- | --- |
| Trailmaking Errors | 116.9% | +5.9% to +412.6% | 4.496 |  | **0.0340** |

^1^ Beta coefficients represent the estimate percent difference in trailmaking errors associated with
status

**Supplemental table 6. Summary statistics for using MBI-C status (Cutpoint of 6) to predict Lumosity tasks (Ordinal Logistic Regression)**

| Outcome | Odds Ratio | 95% CI | *p* value |
| --- | --- | --- | --- |
| GoNoGo Errors | 0.987 | 0.496 to 1.919 | 0.969 |
